# Diffusion tensor imaging analysis along the perivascular space suggests impaired glymphatic clearance in Lewy body dementia subtypes

**DOI:** 10.1101/2025.11.21.25340741

**Authors:** Naomi Hannaway, Angeliki Zarkali, Rohan Bhome, Ivelina Dobreva, Sabrina Kalam, George EC Thomas, Barbara Dymerska, Irene Gorostiaga Belio, Katie Tucker, Amanda Heslegrave, Henrik Zetterberg, Rimona S Weil

## Abstract

**Background:** Parkinson’s disease dementia (PDD) and dementia with Lewy bodies (DLB) are subtypes of Lewy body dementia (LBD) and overlap in symptoms and pathology. Glymphatic function is implicated in LBD pathophysiology due to reduced clearance of abnormal proteins. We aimed to investigate differences in diffusion tensor imaging along the perivascular space (DTI-ALPS), a candidate *in vivo* measure of glymphatic function, between LBD sub-types. We compared DTI-ALPS between DLB, PDD, Parkinson’s with normal cognition (PD-NC), and control participants.

**Methods:** We recruited participants with DLB, PDD, PD-NC, and controls from neurology clinics and patient support groups. Participants were aged 50-80, clinically diagnosed. Exclusions were confounding neurological or psychiatric conditions or metal precluding MRI scanning. Participants underwent MRI brain, plasma sampling and clinical and cognitive assessments. DTI-ALPS was calculated for each participant. As DTI-ALPS can be influenced by white matter distribution, we also calculated a metric called “complexity”. We tested group differences in DTI-ALPS, and associations with clinical variables across all patient groups including cognition, motor scores, sleep and fluctuations.

**Results:** Fifty-one DLB (43 M, 72.7±5.5 years), 35 PDD (25 M, 69.5±7.8 years), 60 PD-NC (27 M, 63.1±7.33 years), and 26 controls (13 M, 66.7±9.28 years), were included for analysis. DTI-ALPS significantly differed between groups (F(3, 165) = 22.68, p<.0001), controlling for age and sex. DTI-ALPS did not differ between PD-NC and controls, whilst cognitively impaired groups (PDD, DLB) had reduced DTI-ALPS relative to PD-NC and to controls(p_FDR_<.05). DTI-ALPS was further reduced in DLB relative to PDD. These findings held when corrected for complexity, which accounts for differences in white matter distribution (F(3, 161) = 28.11, p<.0001). Finally, DTI-ALPS correlated with cognition (MOCA: β=2.95, p<.0001; composite cognitive score: β=1.81, p=.046), REM sleep behaviour disorder (β=-4.36, p=.019), and cognitive fluctuations (β=14.89, p=.014).

**Conclusions:** We showed that DTI-ALPS, is reduced in LBD compared to PD-NC and controls; and is further reduced in DLB compared to PDD. DTI-ALPS is an easily-extracted metric from widely-used MRI scans which has potential as a useful imaging marker in LBD for clinical trials and in the clinical setting, to identify LBD patients with a more aggressive disease course.

## Introduction

Dementia with Lewy bodies (DLB) and Parkinson’s disease dementia (PDD) are characterised by dementia accompanied by core clinical features of motor parkinsonism alongside visual hallucinations, cognitive fluctuations and rapid eye movement (REM) sleep behaviour disorder.^1^ Together, they are referred to under the umbrella term Lewy body dementia (LBD), the second most common cause of degenerative dementia after Alzheimer’s disease.^2^ The distinction between DLB and PDD is based on the timings of dementia onset: DLB is diagnosed when dementia occurs prior to or within one year of motor symptom onset, whereas PDD is diagnosed when dementia occurs over a year after onset of motor parkinsonism. There is continued debate whether DLB and PDD are part of a single disease spectrum^3^ or are two separable diseases^4^, but recent research using advanced imaging techniques suggests that there are measurable differences between these groups, including differences in white matter integrity^5,6^.

LBD is characterised by a combination of pathologies. In addition to α-synuclein pathology, including Lewy bodies and Lewy neurites, the majority of patients with LBD show beta-amyloid and tau pathology in the brain^7,8^, with greater beta-amyloid deposition found in DLB than in PDD^7,9^. Together, these pathological accumlations are thought to drive dementia progession and influence motor and cognitive symptomology of LBD^10^.

The clearance of waste and extracellular proteins, including a-synuclein, amyloid and tau, is proposed to occur through the glymphatic system^11^; with glymphatic clearance recently implicated in the pathophysiology of neurodegenerative disease due to impaired removal of pathological accumulations^12^. Glymphatic clearance is thought to occur through convective flow of interstitial fluid, facilitated by aquaporin-4 channels within astrocytes^11^. *In vivo* techniques have recently been developed, allowing measurement of the glymphatic system, but require intrathecal injection of tracers and sequential imaging^13^. Diffusion tensor image analysis along the perivascular space (DTI-ALPS) measures the diffusion of water molecules in the perivascular space at the level of the body of the lateral ventricle^14^. It correlates strongly with the gold standard method for measuring glymphatic function using intrathecal injection of contrast and sequential MRI imaging^15^, but offers a non-invasive and more widely accessible alternative^16^. Furthermore, DTI-ALPS correlates with clinical metrics known to be associated with glymphatic clearance including sleep quality, age, and cognitive performance^17^.

Several studies have examined DTI-ALPS in PD ^18–24^ and in LBD ^25–28^, showing reduced DTI-ALPS in PD and in LBD compared with controls, associations with poorer cognition in PD and LBD ^21,25,27^, and worse outcomes in PD^29^. Similarly, DTI-ALPS is decreased in and Alzheimer’s disease (AD)^30^, where it is also associated with worse cognitive scores^30^ and with higher tracer uptake on amyloid- and tau-PET^15^. However, there have been no studies comparing DTI-ALPS between patients with LBD and PD with normal cognition, or between DLB and PDD.

Given that postmortem evidence suggests greater amyloid deposition in DLB than PDD^7,9^, and that the glymphatic system is implicated in the removal of pathological proteins including beta-amyloid from the brain, measurements of glymphatic function may be sensitive to differences between PDD and DLB groups in life. Here, we investigated whether DTI-ALPS differs between patients with LBD and PD with normal cognition, between subtypes of LBD (PDD and DLB), and whether it relates to clinically relevant measures in LBD.

First, we aimed to test whether DTI-ALPS differed between LBD and people with PD who had normal cognition (PD-NC) or controls; and between PDD and DLB. Next, we aimed to ensure that any differences seen were not due to variations in white matter integrity and tested this by applying a metric called complexity. Finally, we tested whether DTI-ALPS was associated with core clinical symptoms of LBD, and with other disease-relevant measures including plasma biomarkers and small vessel disease. We predicted reduced DTI-ALPS in PDD and DLB relative to PD-NC and controls. Based on previous postmortem work reporting greater beta-amyloid deposition in DLB, we also predicted reduced DTI-ALPS in DLB relative to PDD. We expected that this would survive correction for complexity; and that lower DTI-ALPS would relate to poorer cognitive and motor symptoms in LBD.

## Materials and Methods

### Participants

Participants were recruited from the National Hospital for Neurology and Neurosurgery and affiliated hospitals, the existing Vision in Parkinson’s cohort study which has been previously described^31^, and patient support groups (Rare Dementia Support and Lewy Body Society).

People with DLB and PDD met criteria for a clinical diagnosis according to Diamond Lewy toolkits for DLB or PDD^32^. People with PD-NC met the Movement Disorders Society (MDS) clinical diagnostic criteria for PD^33^. All patients were within 10 years of diagnosis. For PDD, patients were within 10 years of the diagnosis of dementia. Participants with a history of confounding neurological or psychiatric disorders, metal implants considered unsafe for MRI, or atypical Parkinsonism were excluded. Controls diagnosed with dementia or mild cognitive impairment (MCI), or with a Mini Mental State Examination (MMSE) score of less than 25 were also excluded^34^. We also included patients with diagnoses of PD-MCI and MCI-LB, according to MDS and McKeith criteria^35,36^. As numbers were low in these groups, we combined them with the PDD and DLB patients respectively but have also examined MCI separately (see Supplemental Results). It was not possible to separately examine MCI-LB and PD-MCI due to low numbers. All participants gave written informed consent, and the study was approved by the Queen Square Research Ethics Committee (15.LO.0476).

### Clinical and neuropsychological evaluation

All participants underwent clinical and neuropsychological assessments, including the MMSE and Montreal Cognitive Assessment (MoCA), as measures of global cognition, plus two tests per cognitive domain (Attention: Stroop colour naming and digit span; Executive functions: category fluency^37^ and Stroop interference Language: letter fluency and graded naming task; Memory: word recognition task and logical memory; Visuospatial function: Hooper visual organization test and Benton Judgement of line orientation).

Where it was not possible to complete the full neuropsychological battery in patients with LBD due to patient fatigue, the tasks were prioritised in the following order:

1. MMSE and MoCA: all participants completed MoCA; MMSE missing for 1 LBD participant
2. One task from each cognitive domain: namely Stroop colour naming (attention), category fluency (executive functions), letter fluency (language) and Hooper visual organization test (visuospatial function). One or more tasks were missing for 14 LBD participants.
3. All remaining neuropsychology tasks. One or more tasks were missing from most LBD participants; only 16 were able to complete all tasks.

Additionally, if participants were unable to complete the full Stroop task, they completed a ‘Half-Stroop’ or ‘Victoria Stroop’ consisting of the first three lines of the task^38^. Three LBD participants completed a Half-Stroop for the colour naming condition. 57 LBD participants completed a Half-Stroop for the interference condition.

Motor symptom severity was measured (whilst participants were on their usual medication) using the United Parkinson’s Disease Rating Scale, part 3 (UPDRS-III)^39^. Total PD symptoms were measured using the sum of the UPDRS parts I-IV. Cognitive fluctuations were measured in LBD patients using the clinician assessment of fluctuations (CAF), one-day fluctuations scale^40^, and the dementia cognitive fluctuation scale (DCFS)^41^. Anxiety and depression were measured using the Hospital Anxiety and Depression Scale (HADS)^42^ or the Generalised Anxiety Disorder questionnaire (GAD-7)^43^ and Patient Health Questionnaire (PHQ-9)^44^.

Impairments in activities of daily living were measured using the Functional Activities Questionnaire;^45^ autonomic symptoms using Compass-31;^46^ sleep disturbances using the Rapid Eye Movement Behaviour Disorder Sleep Questionnaire (RBDSQ)^47^ and visual hallucinations using the University of Miami PD hallucinations questionnaire (UMPDHQ)^48^. Participants reported on vascular risk factors, namely: history of angina, myocardial infarction, stroke, diagnosis of diabetes, high cholesterol, high blood pressure, and smoking status. Similar to previous studies, a total score for history of vascular risk factors, ranging from 0 to 7, was calculated^49,50^. Participants completed all assessments whilst taking their usual medications and Levodopa Equivalent Daily Dose (LEDD) was calculated^51^.

A composite cognitive score was derived using the z-scored average of the MOCA plus one task from each cognitive domain (prioritised in part 2 above). In our cohort, the NPI-4 was not available, however, we derived an estimate of the NPI-4, using clinical measures which were available (HADS or PHQ-9, UMPDHQ and UPDRS-I; for further details, see Supplementary Methods, Supplementary Table 1). A composite LBD symptom score was derived using the averaged NPI-4 estimate, UPDRS-III, DCFS and MOCA^52^.

### Plasma collection and processing

Approximately 30ml of blood was collected in polypropylene EDTA tubes. Samples were centrifuged at 2000g for 10 minutes, generating up to 16×0.5ml plasma and 14×0.5 ml serum aliquots, and stored immediately at −80°C.

P-tau217 concentration was measured using the ALZpath Simoa HD-X p-tau217 Advantage-PLUS kit^53^. 143 participants had a sample available for inclusion in p-tau217 analyses (39 DLB, 31 PDD, 52 PD-NC and 21 controls). NfL and GFAP concentrations were measured using the Simoa Human Neurology 4-Plex A (N4PA) assay (Quanterix). For 38 participants, concentrations of NfL were measured using the Simoa NF-Light assay (Quanterix), and no GFAP measurement was available. 118 participants had a sample available for inclusion in NfL analysis (28 DLB, 29 PDD, 47 PD-NC and 14 controls). 78 participants had a sample available for inclusion in GFAP analysis (23 DLB, 18 PDD, 23 PD-NC and 14 controls).

Plasma measurements were performed by analysts blinded to diagnoses and clinical data. P-tau217 measurements were performed in a single batch of reagents, and NfL and GFAP (4-Plex) measurements were performed in 4 batches of reagents. There was no effect of batch on NfL or GFAP concentrations.

### MRI acquisition

All MRI data was acquired on the same 3T Siemens Prisma scanner (Siemens) with a 64-channel coil. The following parameters were used for acquisition: T1-weighted magnetisation-prepared rapid gradient-echo (MPRAGE) images - 1×1×1mm voxel, matrix dimensions 256×256×176, echo time (TE)=3.34ms, repetition time (TR)=2530ms, inversion time(TI)=1100ms, flip angle=7 degrees; diffusion weighted imaging (DWI): 2×2×2mm isotropic voxels, matrix dimensions 220×220×72, TE=58ms, TR=3260ms, *b*=50s/mm^2^/17 directions, *b*=300s/mm^2^/8 directions, *b*=1000s/mm^2^/64 directions, b=2000s/mm^2^/64 directions, and in-plane acceleration factor=2 and multi-band acceleration factor=2; fluid attenuated inversion recovery (FLAIR): 1×1×1mm voxels, matrix dimensions 256×256×192, TE=403ms, TR=4800ms; TI=1650ms, T2 weighted imaging: 1×1×1mm voxels, matrix dimensions 256×256×192, TE=404ms, TR=3200ms, TR=25ms; susceptibility-weighted imaging (SWI): 3D flow-compensated spoiled-gradient-recalled echo sequence, 1×1×1mm voxels, matrix dimensions 204×224×160, TE=18ms, flip angle=12 degrees, receiver bandwidth=110Hz/pixel. For 8 participants, multi-echo fast low angle shot (FLASH) scans with proton density-weighting were acquired – TR=25ms, TE1=2.3ms and TE8=18.4ms (equidistant echoes), flip angle=6 degrees, bandwidth=488Hz/pixel, matrix dimensions=224×256×180 with 1×1×1mm voxels. FLASH magnitude and phase were reconstructed with the MORSE framework^54^ and then used to generate SWI images via the CLEAR-SWI pipeline, as described previously^55^ and detailed in the supplementary methods.

### Image Analysis

#### DTI-ALPS

Diffusion MRI images were pre-processed using Mrtrix3 by denoising,^56^ removal of Gibbs ringing artefacts,^57^ eddy current correction, motion correction^58^ and bias-field correction^59^. Diffusion-weighted images were up-sampled to 1.3mm isotropic resolution. The diffusion tensor was calculated from the pre-processed diffusion images. Fibre orientation distributions (FODs) were computed using multi-shell 3-tissue-constrained spherical deconvolution using the group-average response function for each tissue type (grey matter, white matter & cerebrospinal fluid)^60,61^. Each participant’s FOD image was registered to a study specific template using affine and then nonlinear registration, and a template mask image calculated as the intersection of the brain masks for each participant.

As described previously^29^, DTI-ALPS was calculated in Mrtrix3 using custom scrips, based on Taoka et al^17^. Briefly, diffusivity maps were generated along the x-(Dxx), y-(Dyy) and z (Dzz) axes. An automated atlas-based approach was used to identify association and projection fibres^16^. ROIs were defined based on the John Hopkins University atlas for the association fibres/superior longitudinal fasciculus (SLF; centre co-ordinates left −128,110,99, right - 51,110,99) and projection fibres/superior corona radiata (SCR; left-116,110,99; right-64,110,99).

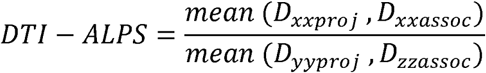

DTI-ALPS was calculated separately for the right and left hemispheres. There was no difference in DTI-ALPS between the right and left hemispheres (t=1.02, p=.31). Therefore, the mean of both hemispheres was calculated and used for all subsequent analyses.

As DTI-ALPS can be influenced by the white matter distribution within calculated ROIs, we used a “complexity” metric. This reflects the difference in orientation of different fibre bundle populations within each voxel and their relative sizes, and can be used to correct for these differences^62^. Complexity ranges in value from zero to one; with values close to zero indicating voxels where all fibres have the same orientation (a single fibre bundle) and higher values indicating voxels where the largest fibre bundle within a voxel contains fewer of the total number of fibres, i.e. when the structure is more complex.^62^ Whilst conventional measures derived from diffusion tensor models, such as fractional anisotropy or mean diffusivity, have limited sensitivity to regions with crossing fibres, this can usually be overcome by using a higher-order diffusion model^63^. DTI-ALPS is derived from the diffusion tensor model, therefore, the complexity metric, which is derived from a fixel-based computational model is used here to provide additional information on crossing fibres and account for these. The complexity metric was calculated for each ROI within each subject using Mrtrix3 as follows: a voxel-wise measure of complexity was calculated across the whole brain using the complexity operation within the fixel2voxel command in mrtrix3^62^.

This approximates FOD peaks by using Bingham distributions, comparing the fibre density of different peaks within a voxel to provide more information on the white matter microstructure^62^. Complexity was averaged across the SLF and SCR ROIs in each hemisphere to give an average complexity within each ROI where DTI-ALPS was calculated for each participant.

### Small Vessel Disease

As DTI-ALPS is influenced by white matter integrity, which is also affected by vascular change, we aimed to quantify small vessel disease in each participant. We used established visual rating scales to quantify the four major forms of cerebral small vessel disease as follows. Briefly:

1. White matter hyperintensities (WMH) were rated on FLAIR images using the modified Fazekas scale^64^.
2. Cerebral microbleeds (CMB): were rated on SWI images using the validated MARS visual rating scale to count and record CMB locations^65^.
3. Lacunes of presumed vascular origin were visually identified on FLAIR images as hypointense lesions, between 3-15mm in diameter, following STRIVE-2 criteria^66^.
4. Enlarged perivascular spaces (EPVS) were rated in basal ganglia on T2-weighted MRI following a standardised visual rating scale^67^.

Each of these measures were scored by 2 raters who were blind to the DTI-ALPS score and to patient outcomes at the time of scoring (ID, SK), and any discrepancies resolved by a third rater who was also blind to these aspects (AZ). Across all measures, inter-rater agreement percentage was 89%. A total SVD burden score ranging from 0 to 4 was calculated according to STRIVE criteria^68^ with a point awarded for each of the following MRI features of SVD: presence of at least one lacune, presence of at least one CMB, moderate to severe (grade 2–4)

EPVS in the basal ganglia, and modified Fazekas score of 2 or 3^69^. See Supplementary Methods for more details on the scoring and agreement ratings of each feature.

### Statistical analysis

Comparisons were made between disease groups using one-way analysis of variance (ANOVA) tests, controlling for age and sex. Planned comparisons were conducted to compare each disease group to controls (a. controls vs PD-NC, b. controls vs PDD, c. controls vs DLB), each LBD group to PD-NC (d. PD-NC vs PDD, e. PD-NC vs DLB) and f. PDD vs DLB.

Associations between DTI-ALPS and cognitive/clinical scores were tested across combined patient groups (DLB, PDD, PD), using single linear regression with the variable of interest, and using multiple linear regression including age as a covariate and including age, sex, and average complexity as covariates. P-values were adjusted for multiple comparisons (6 pairwise comparisons for group analyses), using false discovery rate and p _FDR_<.05 was accepted as the threshold for statistical significance. Statistical analyses were performed in R (R-4.2.1; https://www.r-project.org/).

### Standard Protocol Approvals, Registrations, and Patient Consents

All participants gave written informed consent, and the study was approved by the Queen Square Research Ethics Committee (15.LO.0476).

## Results

### Demographics and Clinical Severity

A total of 51 people with DLB, 35 PDD, 60 PD-NC, and 26 age-matched controls were included for analysis. We grouped PD-MCI together with PDD, and LB-MCI together with DLB, but also examined MCI separately (see supplemental Results for details). Patients with DLB and PDD were older than those with PD-NC, and DLB patients were older than controls. Both DLB and PDD had a greater proportion of males compared to PD-NC, and DLB had a greater proportion of males than controls. As expected, patients with DLB and PDD had lower cognitive scores (MMSE, MoCA, composite cognitive score) than controls and PD-NC. UPDRS-III and total UPDRS scores were greater for all patient groups (DLB, PDD, PD-NC) than controls and were higher for DLB and PDD compared to PD-NC (Table 1 and Table 2).

**Table 1.** Demographic and clinical information of participants.

|  | Controls<br>(n = 26) | PD<br>(n = 60) | PDD<br>(n = 35) | DLB<br>(n = 51) | Test statistic | p-value |
| --- | --- | --- | --- | --- | --- | --- |
| Age | 66.7 (9.28) | 63.1 (7.33) <sup>a</sup> | 69.5 (7.8) <sup>b</sup> | 72.7 (5.5) <sup>a, b, c</sup> | $\chi^2 = 42.07$ | **<.0001** |
| Gender | 13 M / 13 F | 27 M / 33 F | 25 M / 10F <sup>b</sup> | 43 M / 8 F <sup>a, b</sup> | $\chi^2 = 19.6$ | **<.0001** |
| Years Education | 17.4 (2.9) | 16.3 (2.6) | 15.3 (4.0) | 14.8 (3.2) | $\chi^2 = 0.2$ | .62 |
| UPDRS-III | 5.65 (4.8) | 19.9 (10.1) <sup>a</sup> | 30.6 (13.2) <sup>a, b</sup> | 36.4 (18.0) <sup>a, b</sup> | $\chi^2 = 72.4$ | **<.0001** |
| UPDRS-total | 9.3 (5.9) | 40.3 (18.7) <sup>a</sup> | 57.0 (27.3) <sup>a, b</sup> | 71.7 (29.5) <sup>a, b, c</sup> | $\chi^2 = 88.3$ | **<.0001** |
| LEDD | - | 473 (282) | 750 (386) | 285 (267) | $\chi^2 = 38.1$ | **<.0001** |
| Disease Duration<br>Parkinsonism | - | 5.9 (14.2) | 8.2 (5.4) | 2.4 (2.1) | $\chi^2 = 44.0$ | **<.0001** |
| Disease Duration<br>Cognition | - | - | 1.8 (2.2) | 2.4 (2.1) | W = 538.5 | **.002** |
| Global cognitive scores |  |  |  |  |  |  |
| MMSE | 29.1 (1.0) | 29.3 (0.8) | 27.7 (2.5) <sup>a, b</sup> | 23.5 (4.0) <sup>a, b, c</sup> | $\chi^2 = 80.6$ | **<.0001** |
| MOCA | 28.8 (1.3) | 28.7 (1.2) | 24.5 (4.2) <sup>a, b</sup> | 20.6 (5.2) <sup>a, b, c</sup> | $\chi^2 = 102.9$ | **<.0001** |
| Composite Cognitive<br>Score | 0.02 (0.6) | -0.01 (0.6) | -1.34 (1.3) <sup>a, b</sup> | -2.84 (1.8) <sup>a, b, c</sup> | $\chi^2 = 85.5$ | **<.0001** |
| **Plasma measures** |  |  |  |  |  |  |
| p-tau217 | 0.43 (0.32) | 0.36 (0.17) | 0.46 (0.26) | 0.58 (0.29)<sup>a, b</sup> | $\chi^2 = 17.1$ | **.0005** |
| NfL | 16.28 (9.87) | 14.84 (5.69) | 18.9 (7.37)<sup>b</sup> | 23.1 (11.63)<sup>a, b</sup> | $\chi^2 = 16.6$ | **.0009** |
| GFAP | 137.4 (73.4) | 134.7 (83.9) | 150.3 (79.7) | 188.5 (110.2) | $\chi^2 = 4.5$ | .21 |
| **Measures of Small Vessel disease** |  |  |  |  |  |  |
| History of vascular risk factors | 0.71 (0.83) | 0.60 (0.90) | 0.79 (0.89) | 0.75 (0.99) | $\chi^2 = 2.8$ | .42 |
| Total SVD score+ | - | - | 2.25 (0.96) | 1.89 (0.81) | W = 47 | .47 |

All values are mean(standard deviation), apart from gender which is shown as a proportion; DLB= Dementia with Lewy bodies, GFAP = glial fibrillary acidic protein, LEDD = Levodopa equivalent daily dose, MMSE = Mini Mental State Examination, MoCA = Montreal Cognitive Assessment, NfL = neurofilament light chain, PD = Parkinson's disease; PDD = Parkinson's Dementia; p-tau217 = tau phosphorylated at threonine 217, SVD = small vessel disease, UPDRS-III = Unified Parkinson's Disease Rating Scale part 3 (motor assessment). For people with PDD, disease duration is specified separately for onset of motor Parkinsonism and onset of dementia. Participants with PD-MCI and MCI-LB were grouped with PDD and DLB groups respectively as numbers were low in these groups. Significant differences between groups were examined with post-hoc Dunn test. These are displayed as <sup>a</sup> Significantly different from controls (for all other groups). <sup>b</sup> significantly different from PD (PDD and DLB). <sup>c</sup> significantly different from PDD (for DLB only)

**Table 2.**
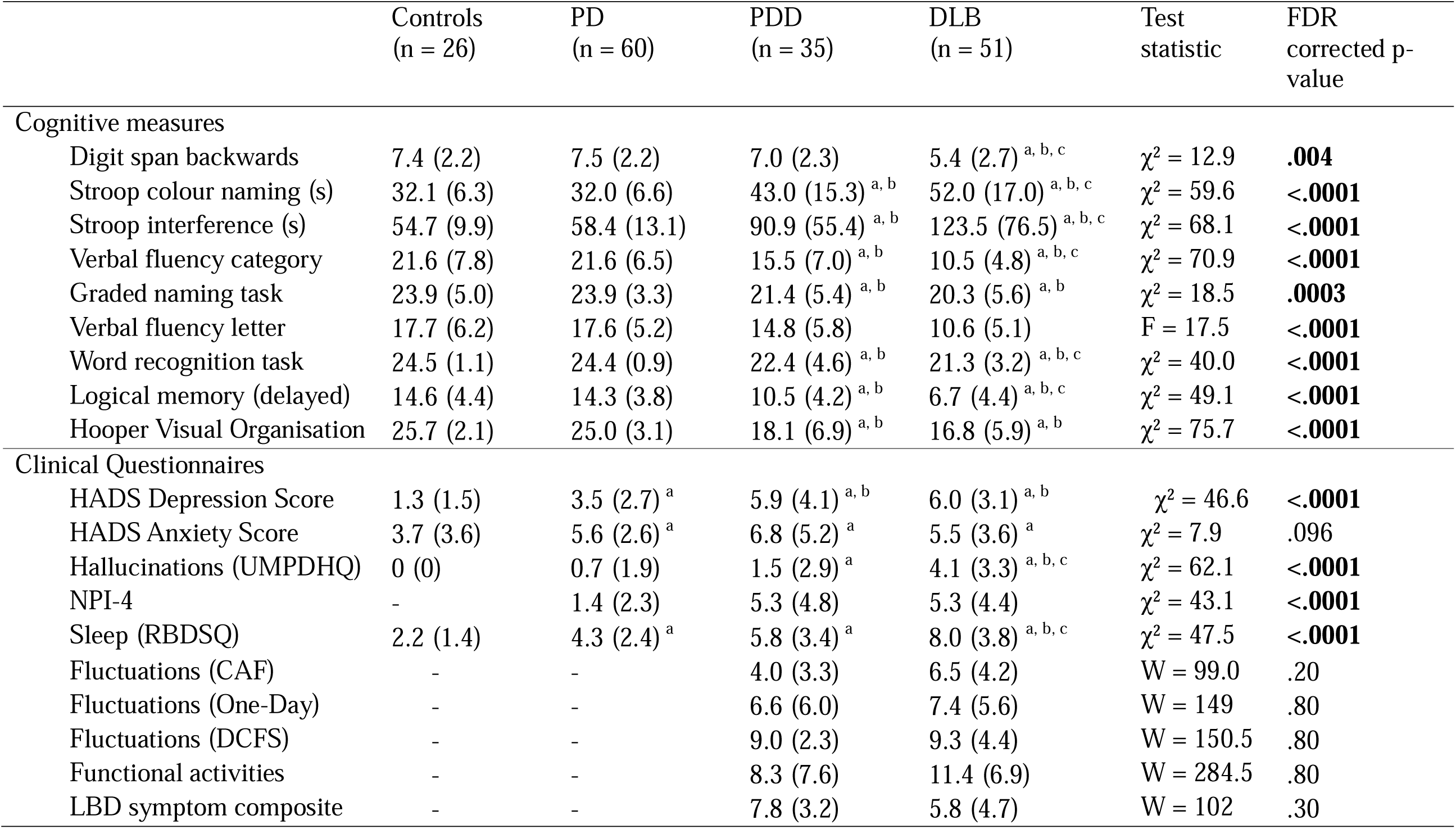
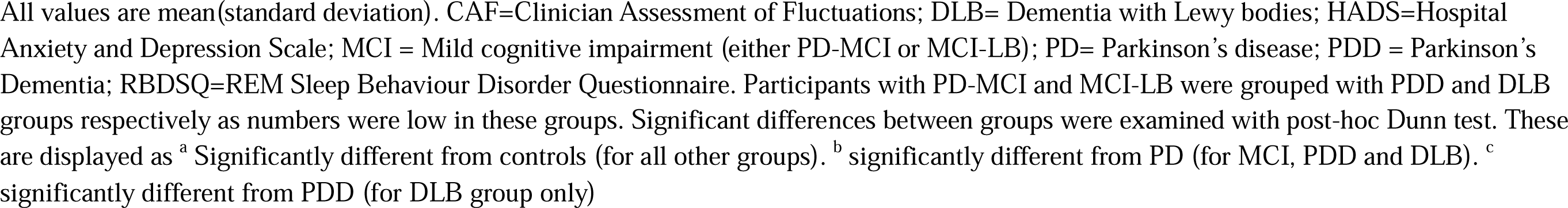
Cognitive and questionnaire scores.

|  | Controls<br>(n = 26) | PD<br>(n = 60) | PDD<br>(n = 35) | DLB<br>(n = 51) | Test<br>statistic | FDR<br>corrected p-value |
| --- | --- | --- | --- | --- | --- | --- |
| **Cognitive measures** |  |  |  |  |  |  |
| Digit span backwards | 7.4 (2.2) | 7.5 (2.2) | 7.0 (2.3) | 5.4 (2.7)<sup>a, b, c</sup> | $\chi^2 = 12.9$ | **.004** |
| Stroop colour naming (s) | 32.1 (6.3) | 32.0 (6.6) | 43.0 (15.3)<sup>a, b</sup> | 52.0 (17.0)<sup>a, b, c</sup> | $\chi^2 = 59.6$ | **<.0001** |
| Stroop interference (s) | 54.7 (9.9) | 58.4 (13.1) | 90.9 (55.4)<sup>a, b</sup> | 123.5 (76.5)<sup>a, b, c</sup> | $\chi^2 = 68.1$ | **<.0001** |
| Verbal fluency category | 21.6 (7.8) | 21.6 (6.5) | 15.5 (7.0)<sup>a, b</sup> | 10.5 (4.8)<sup>a, b, c</sup> | $\chi^2 = 70.9$ | **<.0001** |
| Graded naming task | 23.9 (5.0) | 23.9 (3.3) | 21.4 (5.4)<sup>a, b</sup> | 20.3 (5.6)<sup>a, b</sup> | $\chi^2 = 18.5$ | **.0003** |
| Verbal fluency letter | 17.7 (6.2) | 17.6 (5.2) | 14.8 (5.8) | 10.6 (5.1) | F = 17.5 | **<.0001** |
| Word recognition task | 24.5 (1.1) | 24.4 (0.9) | 22.4 (4.6)<sup>a, b</sup> | 21.3 (3.2)<sup>a, b, c</sup> | $\chi^2 = 40.0$ | **<.0001** |
| Logical memory (delayed) | 14.6 (4.4) | 14.3 (3.8) | 10.5 (4.2)<sup>a, b</sup> | 6.7 (4.4)<sup>a, b, c</sup> | $\chi^2 = 49.1$ | **<.0001** |
| Hooper Visual Organisation | 25.7 (2.1) | 25.0 (3.1) | 18.1 (6.9)<sup>a, b</sup> | 16.8 (5.9)<sup>a, b</sup> | $\chi^2 = 75.7$ | **<.0001** |
| **Clinical Questionnaires** |  |  |  |  |  |  |
| HADS Depression Score | 1.3 (1.5) | 3.5 (2.7)<sup>a</sup> | 5.9 (4.1)<sup>a, b</sup> | 6.0 (3.1)<sup>a, b</sup> | $\chi^2 = 46.6$ | **<.0001** |
| HADS Anxiety Score | 3.7 (3.6) | 5.6 (2.6)<sup>a</sup> | 6.8 (5.2)<sup>a</sup> | 5.5 (3.6)<sup>a</sup> | $\chi^2 = 7.9$ | .096 |
| Hallucinations (UMPDHQ) | 0 (0) | 0.7 (1.9) | 1.5 (2.9)<sup>a</sup> | 4.1 (3.3)<sup>a, b, c</sup> | $\chi^2 = 62.1$ | **<.0001** |
| NPI-4 | - | 1.4 (2.3) | 5.3 (4.8) | 5.3 (4.4) | $\chi^2 = 43.1$ | **<.0001** |
| Sleep (RBDSQ) | 2.2 (1.4) | 4.3 (2.4)<sup>a</sup> | 5.8 (3.4)<sup>a</sup> | 8.0 (3.8)<sup>a, b, c</sup> | $\chi^2 = 47.5$ | **<.0001** |
| Fluctuations (CAF) | - | - | 4.0 (3.3) | 6.5 (4.2) | W = 99.0 | .20 |
| Fluctuations (One-Day) | - | - | 6.6 (6.0) | 7.4 (5.6) | W = 149 | .80 |
| Fluctuations (DCFS) | - | - | 9.0 (2.3) | 9.3 (4.4) | W = 150.5 | .80 |
| Functional activities | - | - | 8.3 (7.6) | 11.4 (6.9) | W = 284.5 | .80 |
| LBD symptom composite | - | - | 7.8 (3.2) | 5.8 (4.7) | W = 102 | .30 |

All values are mean(standard deviation). CAF=Clinician Assessment of Fluctuations; DLB= Dementia with Lewy bodies; HADS=Hospital Anxiety and Depression Scale; MCI = Mild cognitive impairment (either PD-MCI or MCI-LB); PD= Parkinson's disease; PDD = Parkinson's Dementia; RBDSQ=REM Sleep Behaviour Disorder Questionnaire. Participants with PD-MCI and MCI-LB were grouped with PDD and DLB groups respectively as numbers were low in these groups. Significant differences between groups were examined with post-hoc Dunn test. These are displayed as <sup>a</sup> Significantly different from controls (for all other groups). <sup>b</sup> significantly different from PD (for MCI, PDD and DLB). <sup>c</sup> significantly different from PDD (for DLB group only)

### DTI-ALPS is lower in DLB and PDD compared with PD-NC

DTI-ALPS differed significantly between groups, controlling for age and sex, F(3, 165) = 22.68, p<.0001, Figure 1. Planned post hoc comparisons showed that DTI-ALPS was reduced in DLB and PDD compared with PD-NC (DLB<PD-NC: F=68.6, p_FDR_<.0001; PDD<PD-NC: F=10.6, p_FDR_=.002); and was reduced in DLB and PDD compared to controls (DLB<control: F=42.2, p_FDR_ <.0001; PDD<control: F=5.6, p_FDR_ =.025);. DTI-ALPS was also reduced in DLB compared to PDD (F=13.6, p_FDR_=.0008).

**Figure 1.**
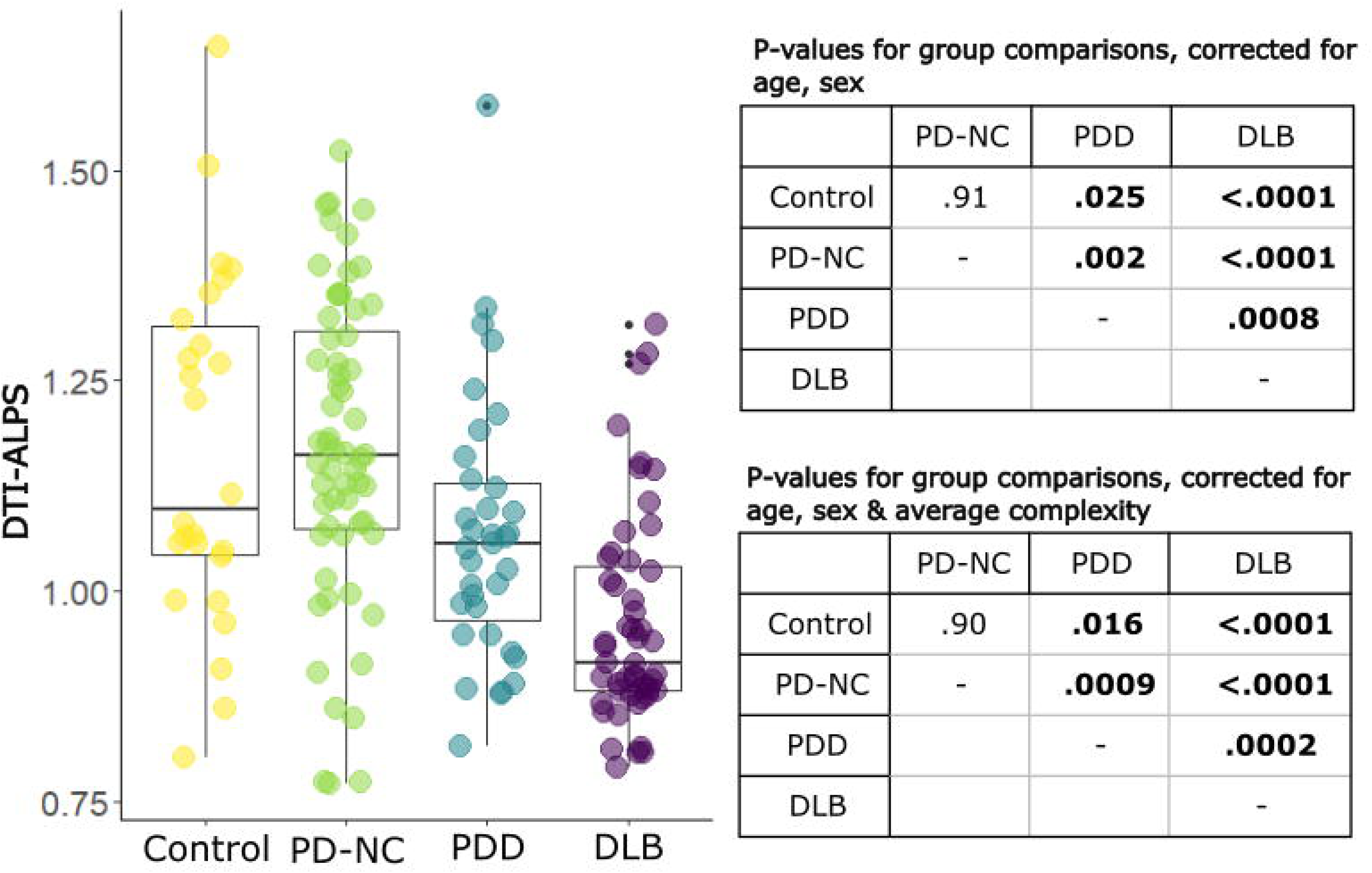
Differences in DTI-ALPS between patients with Parkinson’s disease without cognitive involvement (PD-NC), Parkinson’s disease dementia (PDD) and dementia with Lewy bodies (DLB). DTI-ALPS = diffusion tensor imaging along the perivascular space, PD=Parkinson’s Disease, DLB= dementia with Lewy bodies, PDD=Parkinson’s Disease Dementia. DTI-ALPS uses arbitrary units. Box-and-whisker plots show median, IQR, range, and that black dots represent outliers. Significant p-values are in bold, as assessed by post-hoc pairwise comparisons.

**Figure 2.**
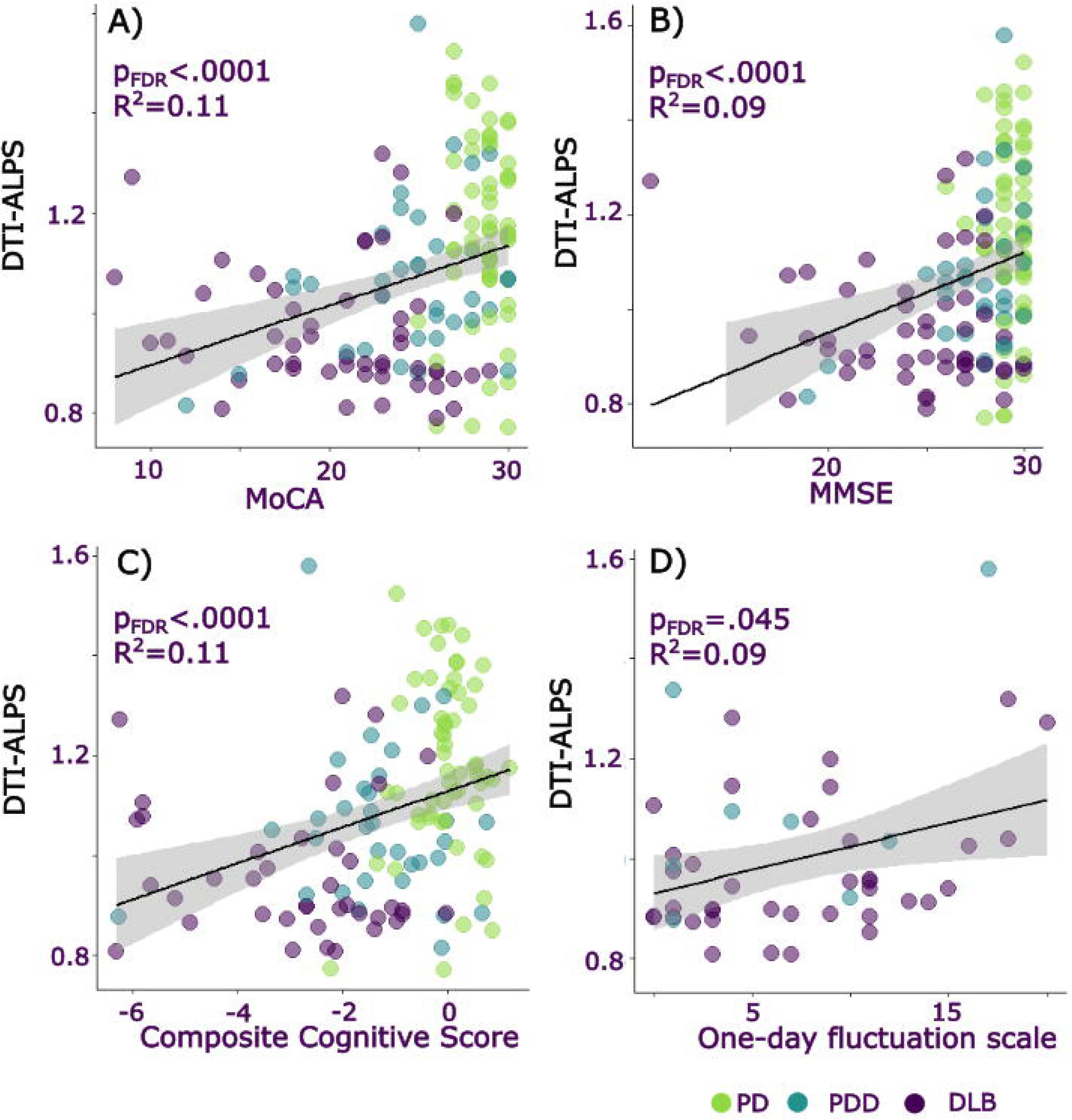
Associations between DTI-ALPS and cognitive measures. Relationship between DTI-ALPS, an indirect measure of glymphatic clearance and A) Montreal cognitive assessment (MoCA), B) Mini-mental state examination (MMSE), C) Composite cognitive score, D) One-day fluctuations scale. Color of dot indicates clinical group: green = PD-NC, blue = PDD, purple = DLB. DLB = Dementia with Lewy Bodies, PD-NC = Parkinson’s disease with normal cognition, PDD = Parkinson’s disease Dementia. DTI-ALPS uses arbitrary units. The raw data for DTI-ALPS and cognitive scores are plotted in the figure; statistical analyses are reported as simple linear regressions and were repeated including age as a covariate and including age, sex and average complexity as covariates. Grey shaded areas represented 95% confidence intervals.

Next, we examined average white matter complexity within the ROIs of DTI-ALPS, to test whether differences in DTI-ALPS were due to differences in white matter distribution. As expected, we found that the complexity metric correlated with DTI-ALPS, β = 0.59, t = 6.80, p<.0001 (Supplementary Figure 1). Average complexity also differed significantly between groups, (F(3, 162) = 12.25, p<.0001, Supplementary Figure 2) and was reduced in DLB and PDD compared to PD-NC (DLB<PD-NC: F=36.9, p_FDR_ <.0001; PDD<PD-NC: F=13.2, p_FDR_ =.0009); was reduced in DLB and PDD compared to controls (DLB<control: F=29.5, p_FDR_ <.0001; PDD<control: F=6.6, p_FDR_ =.019);. There were no differences in average complexity between PDD and DLB (F = 0.0, p_FDR_=.96), nor between PD-NC and controls (F = 0.2, p_FDR_=.66). There were strong correlations between the average complexity within the SCR and SLF ROIs used to calculate DTI-ALPS within each hemisphere (R, r = 0.44, p<.0001; L, r = 0.46, p<.0001; Supplementary Figure 3).

Importantly, when controlling for average complexity, age and sex, DTI-ALPS still differed between groups (F(3, 161) = 28.11, p<.0001), with the same qualitative group differences, when comparisons included a covariate to control for the complexity metric (Figure 1). When we examined MCI groups separately to DLB and PDD groups, the overall pattern of results remained the same (Supplementary Results, Supplementary Figures 4-5).

### DTI-ALPS correlates with clinical severity in LBD

DTI-ALPS was significantly correlated with key clinical measures across all patient groups (Figure 3). Namely, DTI-ALPS showed a positive association with cognition (MMSE, MOCA and composite cognitive score: lower DTI-ALPS relating to poorer cognitive scores). We similarly found a relationship between DTI-ALPS and Parkinsonian motor and overall symptom scores (UPDRS-total, UPDRS-III), plasma markers of NfL and p-tau217, sleep-relevant scales (RBDSQ), One-day fluctuations scale, and history of vascular risk factors (Table 3). DTI-ALPS did not relate to plasma GFAP concentration, autonomic scores (Compass), self-reported sleep quality, other fluctuations scales (CAF, DCFS), small vessel disease scores, nor to a total LBD symptom composite.

**Figure 3.**
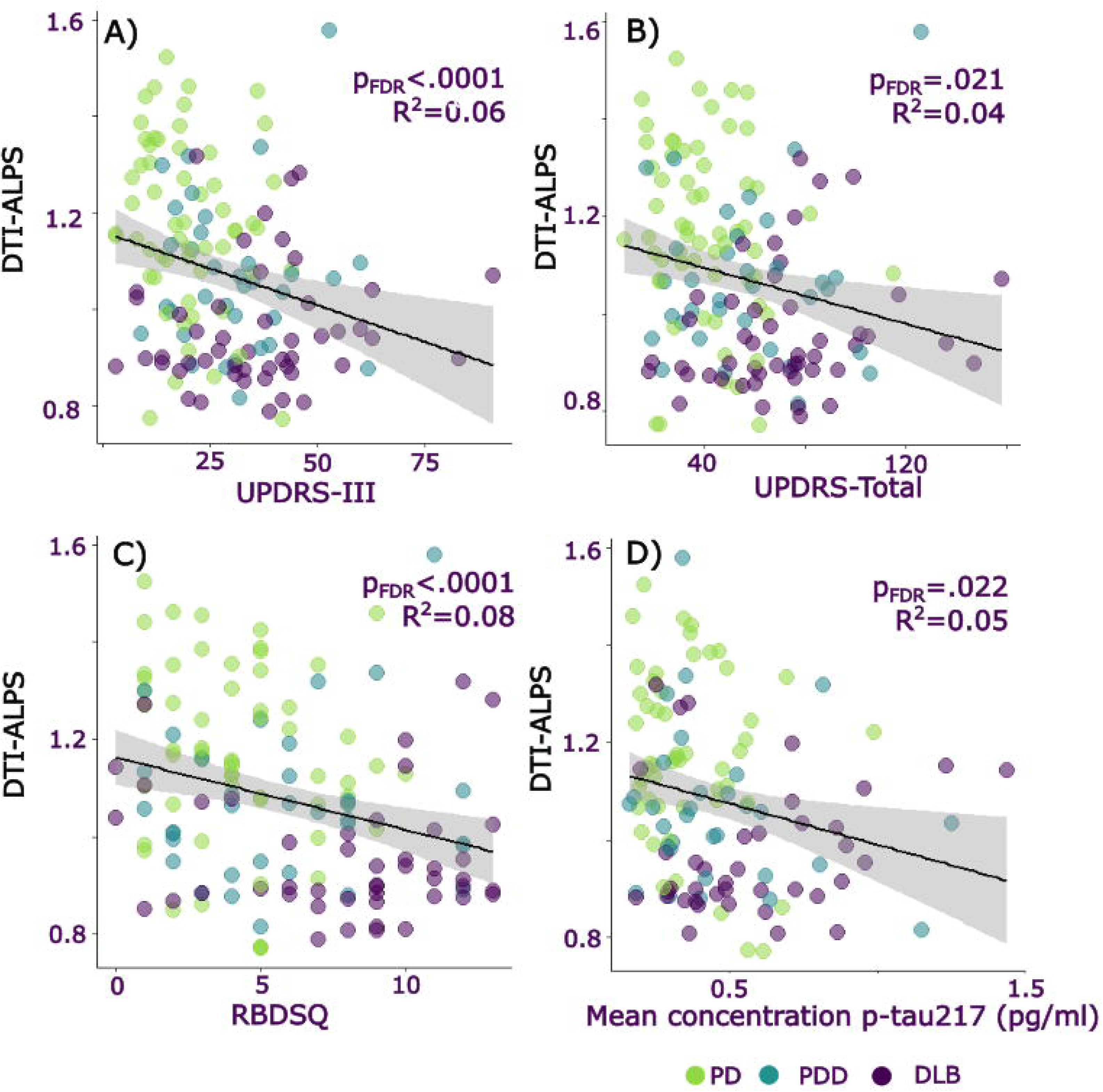
Associations between DTI-ALPS and clinical measures. Relationship between DTI-ALPS, an indirect measure of glymphatic clearance and A) United Parkinson’s disease rating scale – part 3 (UPDRS-III; motor symptom score), B) United Parkinson’s disease rating scale – total symptom score (UPDRS-total), C) Rapid eye movement behaviour disorder sleep quotient (RBDSQ), D) Mean concentration of p-tau217. Color of dot indicates clinical group: green = PD-NC, blue = PDD, purple = DLB. DLB = Dementia with Lewy Bodies, PD-NC = Parkinson’s disease with normal cognition, PDD = Parkinson’s disease Dementia. DTI-ALPS uses arbitrary units. The raw data for DTI-ALPS and cognitive scores are plotted in the figure; statistical analyses are reported as simple linear regressions and were repeated including age as a covariate and including age, sex and average complexity as covariates. Grey shaded areas represented 95% confidence intervals.

**Table 3.**
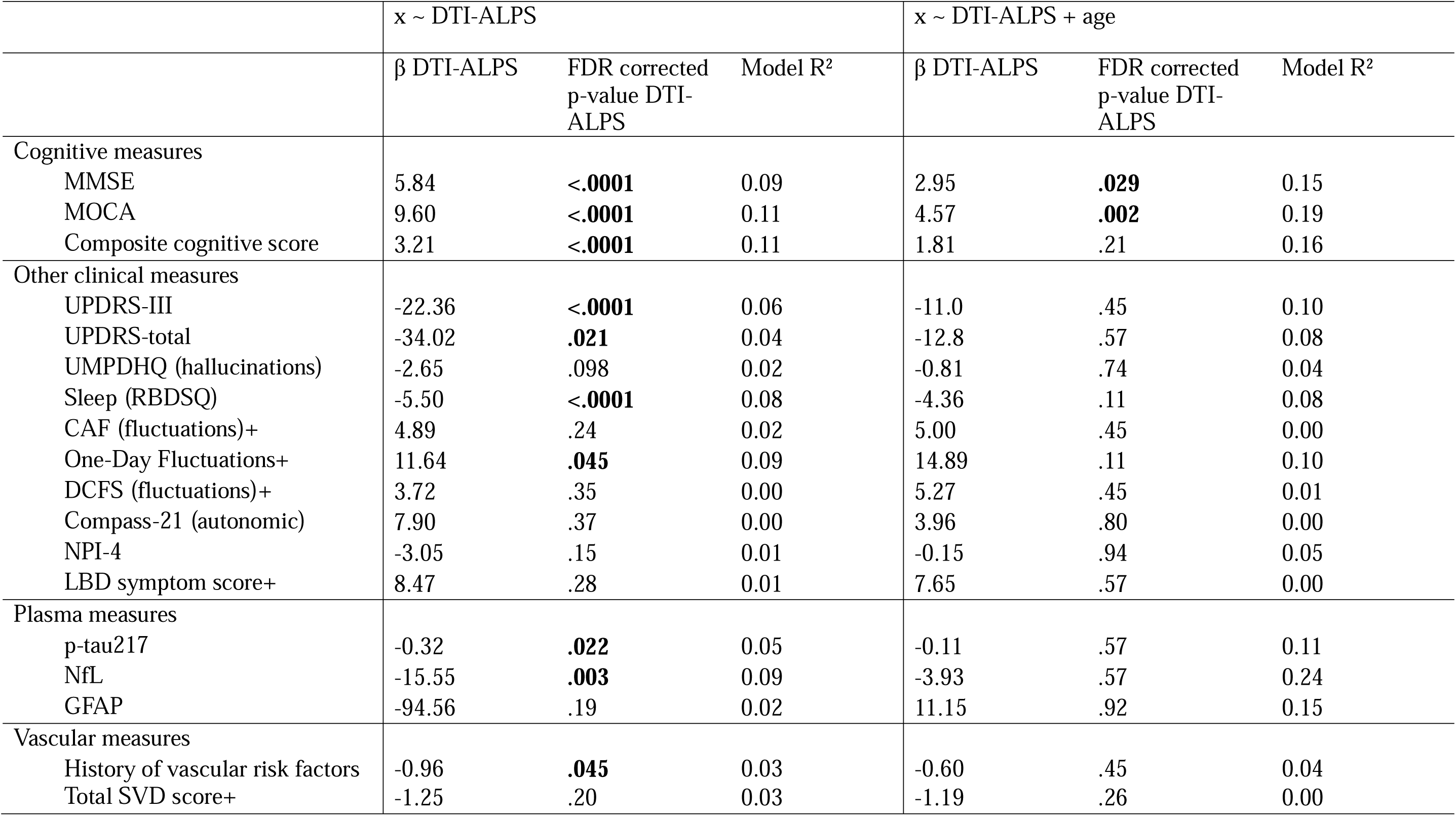
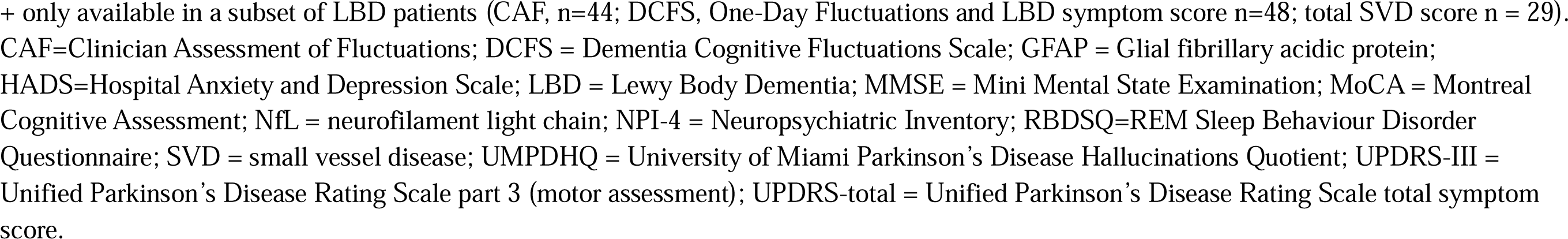
Associations of DTI-ALPS with clinical and cognitive measures; x ∼ DTI-ALPS & x ∼ DTI-ALPS + age.

| | $x \sim \text{DTI-ALPS}$ | | | $x \sim \text{DTI-ALPS} + \text{age}$ | | |
| --- | --- | --- | --- | --- | --- | --- |
| | $\beta$ DTI-ALPS | FDR corrected<br>p-value DTI-ALPS | Model R<sup>2</sup> | $\beta$ DTI-ALPS | FDR corrected<br>p-value DTI-ALPS | Model R<sup>2</sup> |
| Cognitive measures |  |  |  |  |  |  |
| MMSE | 5.84 | **<.0001** | 0.09 | 2.95 | **.029** | 0.15 |
| MOCA | 9.60 | **<.0001** | 0.11 | 4.57 | **.002** | 0.19 |
| Composite cognitive score | 3.21 | **<.0001** | 0.11 | 1.81 | .21 | 0.16 |
| Other clinical measures |  |  |  |  |  |  |
| UPDRS-III | -22.36 | **<.0001** | 0.06 | -11.0 | .45 | 0.10 |
| UPDRS-total | -34.02 | **.021** | 0.04 | -12.8 | .57 | 0.08 |
| UMPDHQ (hallucinations) | -2.65 | .098 | 0.02 | -0.81 | .74 | 0.04 |
| Sleep (RBDSQ) | -5.50 | **<.0001** | 0.08 | -4.36 | .11 | 0.08 |
| CAF (fluctuations)+ | 4.89 | .24 | 0.02 | 5.00 | .45 | 0.00 |
| One-Day Fluctuations+ | 11.64 | **.045** | 0.09 | 14.89 | .11 | 0.10 |
| DCFS (fluctuations)+ | 3.72 | .35 | 0.00 | 5.27 | .45 | 0.01 |
| Compass-21 (autonomic) | 7.90 | .37 | 0.00 | 3.96 | .80 | 0.00 |
| NPI-4 | -3.05 | .15 | 0.01 | -0.15 | .94 | 0.05 |
| LBD symptom score+ | 8.47 | .28 | 0.01 | 7.65 | .57 | 0.00 |
| Plasma measures |  |  |  |  |  |  |
| p-tau217 | -0.32 | **.022** | 0.05 | -0.11 | .57 | 0.11 |
| NfL | -15.55 | **.003** | 0.09 | -3.93 | .57 | 0.24 |
| GFAP | -94.56 | .19 | 0.02 | 11.15 | .92 | 0.15 |
| Vascular measures |  |  |  |  |  |  |
| History of vascular risk factors | -0.96 | **.045** | 0.03 | -0.60 | .45 | 0.04 |
| Total SVD score+ | -1.25 | .20 | 0.03 | -1.19 | .26 | 0.00 |

+ only available in a subset of LBD patients (CAF, n=44; DCFS, One-Day Fluctuations and LBD symptom score n=48; total SVD score n = 29). CAF=Clinician Assessment of Fluctuations; DCFS = Dementia Cognitive Fluctuations Scale; GFAP = Glial fibrillary acidic protein; HADS=Hospital Anxiety and Depression Scale; LBD = Lewy Body Dementia; MMSE = Mini Mental State Examination; MoCA = Montreal Cognitive Assessment; NfL = neurofilament light chain; NPI-4 = Neuropsychiatric Inventory; RBDSQ=REM Sleep Behaviour Disorder Questionnaire; SVD = small vessel disease; UMPDHQ = University of Miami Parkinson's Disease Hallucinations Quotient; UPDRS-III = Unified Parkinson's Disease Rating Scale part 3 (motor assessment); UPDRS-total = Unified Parkinson's Disease Rating Scale total symptom score.

After correcting for age, DTI-ALPS was correlated with MMSE and MoCA only. After correcting for complexity, age and sex, no relationship with DTI-ALPS survived correction for multiple comparisons (See Supplemental Results).

## Discussion

We examined DTI-ALPS, a non-invasive and accessible *in vivo* method that provides an indirect measure of glymphatic clearance, in patients with LBD and PD with normal cognition. We showed that DTI-ALPS is reduced in both DLB and PDD compared with PD-NC, and in DLB compared with controls. It was also reduced in DLB compared to PDD. These group differences remained after correcting for age and for complexity, a measure of white matter distribution. We also found that DTI-ALPS related to key clinical variables in LBD, but after correcting for age, we only found relationships between DTI-ALPS and global cognitive scores.

Our finding of reduced DTI-ALPS in LBD is consistent with other studies in LBD, which have shown reduced DTI-ALPS in DLB^28^, PDD^26,70^ and PD-MCI^26^ relative to controls, but have not examined these patient groups together. We expand upon this by comparing DTI-ALPS between each patient group, showing greater reductions in DTI-ALPS in DLB and PDD compared with PD-NC, and greater reductions in DLB compared to PDD. We found no differences between PD-NC and controls. Although previous studies^18–24^ and meta-analyses^71,72^ report reduced DTI-ALPS in PD relative to controls, most of these appear to have included patients with PD-MCI or PDD, based on reported values for MMSE (24.74 ± 4.71; 26.48 ± 3.97)^18,24^, and MOCA (25.02 ± 2.93; 26.5 ± 0.87-)^20,22^. Where studies explicitly assessed and grouped patients based on cognition, DTI-ALPS in PD-NC did not differ from controls^22^; and patients with PD with cognitive involvement showed reduced DTI-ALPS compared to patients with PD and preserved cognition^21,22^, consistent with our findings.

In prior research, reduced DTI-ALPS has been found to be associated with more severe motor symptoms^71,72^, with poorer cognition in PD ^29,71,72^, LBD^25,27^, and with REM sleep behaviour disorder in PD^22^. Likewise, in this study, we show reduced DTI-ALPS to be related with greater clinical symptom severity; namely with increased UPDRS total and motor symptom scores, decreased cognitive scores, and RBDSQ scores. The most reliable association was with cognitive scores, which remained significant after correcting for age.

DTI-ALPS is thought to reflect glymphatic clearance by measuring water molecule diffusivity in the perivascular space at the level of the body of the lateral ventricle^14^. Although this is an indirect measure, it is strongly associated with gold-standard measures for *in vivo* assessment of glymphatic function^15^. Glymphatic clearance has been suggested to contribute to neurodegeneration across neurodegenerative diseases including PD and LBD, although glymphatic clearance has not yet been directly measured in these conditions. In addition to investigations using DTI-ALPS, this is supported by the postmortem relation between the expression of aquaporin-4 channels and α-synuclein neocortical pathology in PD^73^ and evidence from mouse models showing increased α-synuclein accumulation when glymphatic activity is supressed^74^.

Several mechanisms have been suggested as potential explanations for impaired glymphatic clearance contributing to neurodegeneration in PD and LBD. One model is that reduced glymphatic clearance directly leads to higher levels of pathological protein accumulation. This is supported by findings from animal models showing that both beta-amyloid^75^ and α-synuclein^74^ will accumulate if glymphatic clearance is suppressed. An alternative model is that reduced glymphatic flow allows the spread of misfolded proteins such as abnormal alpha-synuclein, across the extracellular space^12^. It is possible that both these processes contribute to build-up of pathological proteins in LBD^7,8^. Our finding of a reduction of DTI-ALPS, an indirect measure of glymphatic clearance, in DLB compared with PDD, is consistent with results from post-mortem and molecular imaging studies, where pathological protein accumulation is higher in DLB compared with PDD and PD. For example: β-amyloid^76,77^, tau^77^, and α-synuclein^76^ pathology have all been shown to be greater for DLB than PDD and PD. It is possible that reduced glymphatic function may contribute to increases in pathological protein accumulation, although this would need to be tested in future work.

Our finding of reduced DTI-ALPS in DLB compared with PDD also contributes to a developing literature showing that differences between DLB and PDD can be observed in life using a range of advanced neuroimaging techniques which are sensitive to different aspects of tissue composition^5,6^. For example, increased quantitative susceptibility mapping (QSM) values, a metric sensitive to tissue iron, have been shown for PDD versus DLB^6^. It is likely that these approaches are capturing separate information from that detected using DTI-ALPS^29^. By better understanding the disease processes in LBD and where differences are present between DLB and PDD and linking these differences to measures relating to specific protein accumulation, our research could potentially contribute to more tailored treatment approaches in future clinical practice.

We found that whilst DTI-ALPS related to key clinical measures, the majority of these were no longer significant after controlling for age. However notably, cognition remained associated with DTI-ALPS even after correcting for age. This association between DTI-ALPS in LBD and cognitive measures replicates previous work in PD showing that lower DTI-ALPS is associated with poorer cognition^29^.

Despite DTI-ALPS being an indirect measure of clearance, and therefore potentially having a relationship with pathological protein accumulation, we did not find a relationship between DTI-ALPS and plasma concentrations of p-tau217, NfL or GFAP after controlling for age. An association between DTI-ALPS with NfL and GFAP was previously shown by one study in LBD^27^, whilst in PD, NfL and DTI-ALPS were not found to be associated ^29^. Likewise, other recent studies found no association between p-tau181 and DTI-ALPS in PD and LBD^27,29^; nor did they find an association with amyloid-positivity as assessed using amyloid PET^27^.One potential explanation for this lack of association is that pathological protein accumulation in LBD is more complex, with several different proteins accumulating, and the presence of co-pathologies differing between individuals. The accumulation of abnormal α-synuclein pathology may be affected by impaired glymphatic clearance, as well as build-up of beta-amyloid and tau, with a more complex relationship that is not reflected by direct build-up of either protein separately, and differs between individuals. It is also possible that reduced glymphatic clearance is an earlier event; and that pathological accumulation, reflected by higher plasma p-tau217, occurs later, and so was not detected in our patient groups.

Limitations and Future directions There are important methodological limitations of DTI-ALPS as a measure of glymphatic clearance. DTI-ALPS can only be an indirect and deductive measure of glymphatic function. However, it has been shown in previous work to correlate with the gold-standard measurement of intrathecal contrast injection and sequential MRI scanning^15^. It is therefore likely to reflect at least some element of glymphatic function, albeit possibly not fully or solely. Another important consideration is that white matter microstructural differences, including the presence of crossing fibres and axonal undulation have been shown to inflate DTI-ALPS indices^78,79^. To counteract this, we repeated our group analysis and regression analyses including a measure of the average complexity of white matter within the regions of interest used for DTI-ALPS calculation as a covariate. Whilst complexity is not a direct measure of crossing fibres or axonal undulation, a lower complexity would be associated with more accurate derivatives of diffusion in the x and y directions, and therefore a better prediction of the DTI-ALPS. We showed that average complexity was lower in DLB and PDD but did not differ between these groups. Importantly, when we controlled for complexity, this strengthened our group findings of reduced DTI-ALPS in both DLB and PDD relative to PD-NC and controls; and did not qualitatively change the overall pattern of results, suggesting that our findings are relatively robust to the presence of crossing fibres. However, correcting for complexity in our correlational analyses with clinical measures did lead to almost all relationships being lost. This suggests that at least some of these white matter measures and DTI-ALPS are inter-correlated, and white matter integrity rather than DTI-ALPS is likely the primary driver of the correlations with clinical severity.

Our analyses have some limitations. Patients with DLB and PDD differed in age and sex to those with PD-NC. Whilst we corrected our analyses for age and sex, recruitment of matched groups would have been preferable. Participants continued to take their usual dopaminergic medication for MRI scans, to minimise discomfort. However, all comparisons are based on structural imaging and are therefore unlikely to be affected by dopaminergic medication.

Some LBD patients were unable to complete all cognitive tasks, and in some individuals, the half-Stroop was used to estimate a full Stroop and composite cognitive score, which may reduce the reliability of these measures. FLAIR/T2 imaging was available in only a subset of participants, so a smaller sample (n=26) was available for testing the relationship between small vessel disease and DTI-ALPS. Previously, small vessel disease has been shown to be associated with cognition in PD^80,81^ and to DTI-ALPS^15,82^. Whilst we did not find DTI-ALPS to be correlated to small vessel disease scores in this work, this should be validated in a larger cohort with FLAIR/T2 imaging available.

To allow us to test the relationship between DTI-ALPS and psychiatric symptoms relevant to LBD, we approximated the NPI-4 using information available at our cohort. This may reduce the reliability of this measure, and we acknowledge that it would have been preferable to use the standard NPI-4. Our approximation was weakest in detecting delusions, which had a very low incidence in our cohort (1% of patients). The composite LBD symptom score used this NPI-4 approximation, so may also be affected by this limitation.

### Practical implications and significance of this work

Taking into account these limitations and our findings, DTI-ALPs could be a valuable metric in a clinical setting or in clinical trials, as an imaging marker suggesting more severe disease. Whilst DTI-ALPS is not specific to LBD it seems to reflect generally more severe disease and relate to cognitive severity. As such, it could potentially be used to identify more severe patients likely to progress more rapidly, for clinical trials, or to provide prognostic information in the clinic. The DTI-ALPS measure is relatively easy to extract from diffusion imaging scans and has potential to be translated and automated for use in clinical pipelines.

### Summary

In summary, we have shown that patients with PDD and DLB have reduced DTI-ALPS, suggestive of poorer glymphatic clearance, compared with PD-NC, with worse DTI-ALPS in DLB than PDD. Lower DTI-ALPS was also associated with greater cognitive severity. This measure has potential for use in clinical trials and clinical settings to identify patients with more severe disease.

## Data availability

The code for generating DTI-ALPS is available in our previous work^29^. The data that support the findings of this study are available from the corresponding author, upon reasonable request.

## Acknowledgements

We are grateful to the participants in this study who generously and enthusiastically attended for study visits. The authors acknowledge the use of the UCL Myriad High Performance Computing Facility (Myriad@UCL), and associated support services, in the completion of this work.

## Funding

We are grateful to generous support from the Hackett, Harris & Hopkins family. This research was supported by a fellowship from Wellcome to RSW (225263Z/22/Z) and by funding from the Rosetrees trust and UCLH Biomedical Research Centre. AZ is supported by an Alzheimer’s Research UK Clinical Research Fellowship (2018B-001) and receives funding from Rosetrees trust, Academy of Medical Sciences and Parkinson’s UK. RB is supported by a PhD fellowship from Wolfson Foundation and Eisai. BD is supported by the Discovery Research Platform for Naturalistic Neuroimaging, funded by the Wellcome Trust (226793/Z/22/Z). HZ is a Wallenberg Scholar and a Distinguished Professor at the Swedish Research Council supported by grants from the Swedish Research Council (#2023-00356, #2022-01018 and #2019-02397), the European Union’s Horizon Europe research and innovation programme under grant agreement No 101053962, Swedish State Support for Clinical Research (#ALFGBG-71320), the Alzheimer Drug Discovery Foundation (ADDF), USA (#201809-2016862), the AD Strategic Fund and the Alzheimer’s Association (#ADSF-21-831376-C, #ADSF-21-831381-C, #ADSF-21-831377-C, and #ADSF-24-1284328-C), the European Partnership on Metrology, co-financed from the European Union’s Horizon Europe Research and Innovation Programme and by the Participating States (NEuroBioStand, #22HLT07), the Bluefield Project, Cure Alzheimer’s Fund, the Olav Thon Foundation, the Erling-Persson Family Foundation, Familjen Rönströms Stiftelse, Familjen Beiglers Stiftelse, Stiftelsen för Gamla Tjänarinnor, Hjärnfonden, Sweden (#FO2022-0270), the European Union’s Horizon 2020 research and innovation programme under the Marie Skłodowska-Curie grant agreement No 860197 (MIRIADE), the European Union Joint Programme – Neurodegenerative Disease Research (JPND2021-00694), the National Institute for Health and Care Research University College London Hospitals Biomedical Research Centre, the UK Dementia Research Institute at UCL (UKDRI-1003), and an anonymous donor.

## Competing interests

RSW has received speaking and writing honoraria from GE Healthcare, Bial, Omnix Pharma, and Britannia; and consultancy fees from Therakind and Accenture. HZ has served at scientific advisory boards and/or as a consultant for Abbvie, Acumen, Alector, Alzinova, ALZpath, Amylyx, Annexon, Apellis, Artery Therapeutics, AZTherapies, Cognito Therapeutics, CogRx, Denali, Eisai, Enigma, LabCorp, Merck Sharp & Dohme, Merry Life, Nervgen, Novo Nordisk, Optoceutics, Passage Bio, Pinteon Therapeutics, Prothena, Quanterix, Red Abbey Labs, reMYND, Roche, Samumed, ScandiBio Therapeutics AB, Siemens Healthineers, Triplet Therapeutics, and Wave, has given lectures sponsored by Alzecure, BioArctic, Biogen, Cellectricon, Fujirebio, LabCorp, Lilly, Novo Nordisk, Oy Medix Biochemica AB, Roche, and WebMD, is a co-founder of Brain Biomarker Solutions in Gothenburg AB (BBS), which is a part of the GU Ventures Incubator Program, and is a shareholder of MicThera (outside submitted work). All other authors report no competing interests.

## Supplementary material

Supplementary material is available at x.

## Notes

### Author Declarations

Queen Square Research Ethics Committee of University College London gave ethical approval for this work(15.LO.0476).

